# TRACE: A FINE-TUNED BIOMEDICAL LANGUAGE MODEL FOR DIRECTIONALLY INFORMED DRUG REPURPOSING FROM TRANSCRIPTOME-WIDE ASSOCIATION STUDIES

**DOI:** 10.64898/2026.08.25.26361263

**Authors:** Christopher O. Otieno, Hannah M. Seagle, Alexis T. Akerele, James Jaworski, Lindsay Guare, Shefali Setia-Verma, Digna R. Velez Edwards, Todd L. Edwards

**Affiliations:** Division of Epidemiology, Department of Medicine, Vanderbilt Health Nashville, TN 37232, USA; Division of Quantitative and Clinical Science, Department of Obstetrics and Gynecology, Department of Biomedical Informatics, Vanderbilt Health Nashville, TN 37232, USA; Department of Pathology and Laboratory Medicine, University of Pennsylvania Philadelphia, PA 19104, USA; Department of Pathology and Laboratory Medicine, Department of Biostatistics, Epidemiology and Informatics, University of Pennsylvania, Philadelphia, PA 19104, USA

**Keywords:** drug repurposing, transcriptome-wide association study, biomedical language model, natural language processing, precision medicine, endometriosis

## Abstract

Transcriptome-wide association studies (TWAS) can identify genes where genetically predicted gene expression is associated with disease risk, but translating those signals into therapeutic opportunities remains time-consuming, manual, and difficult to reproduce. We developed TRACE (TWAS-driven Repurposing through AI-assisted Curation of Evidence), a gene- and phenotype-agnostic computational pipeline that accepts a TWAS gene and effect-size direction, normalizes the gene symbol, retrieves FDA-approved drug-gene candidates from four online resources, collects related peer-reviewed literature from PubMed, and uses a fine-tuned biomedical language model to classify whether the literature supports a direct drug-gene relationship, the mechanism of action, and the direction of effect. The pipeline then compares the drug-derived direction with the direction implied by the TWAS effect estimate to rank candidate therapeutic pairs and flag potential drug safety concerns. The local classifier, built on BiomedBERT, was trained using pipeline-derived labels, BioCreative VI ChemProt gold-standard chemical-protein relation examples, and author-reviewed active-learning cases, reaching a held-out macro F1 of 0.809 across three simultaneous classification tasks. We validated the pipeline against a manually curated endometriosis gold standard of 43 drug-gene pairs spanning six TWAS-identified genes, developed through S-PrediXcan analysis of endometriosis GWAS summary statistics, manual querying of four drug-gene interaction databases for each gene, literature review of drug-gene mechanistic evidence, and Mendelian randomization validation of candidate pairs. External validation used two independently published genetically informed drug-repurposing studies in metabolic dysfunction-associated steatotic liver disease (MASLD) and type 2 diabetes (T2D). The pipeline recovered 90.7% of endometriosis pairs, 88.2% of MASLD pairs, and 92.9% of T2D pairs that were present in at least one queried database. Applied to 99 endometriosis-associated TWAS genes, the pipeline identified 1,089 FDA-approved drug-gene pairs, 32 candidate therapeutic pairs, and 77 potential safety concerns, including independent recovery of leuprolide acetate, an established endometriosis therapy. This framework provides a scalable, literature-grounded bridge from TWAS discovery to prioritized therapeutic hypotheses, while preserving uncertainty through manual-review flags and requiring downstream Mendelian randomization, electronic health record-based validation, and experimental follow-up before clinical interpretation.

## 1. Introduction

Drug development remains expensive, time-consuming, and failure-prone: recent empirical estimates place the average cost of developing a new drug in the range of hundreds of millions to several billion dollars, and reviews of drug repurposing emphasize the appeal of starting from compounds with established safety, clinical, preclinical, and formulation knowledge that may reduce risk, cost, and development timelines ^1, 2^.

Repurposing approved or investigational drugs can therefore shorten the path from biological insight to testable intervention, but prioritization still requires a credible link between disease biology, drug target, mechanism, and expected direction of effect^2^. Human genetic support improves that link: drug mechanisms with genetic support have been estimated to be 2.6 times more likely to succeed clinically than mechanisms without such support^3^.

Transcriptome-wide association studies (TWAS) methods provide one way to connect genetic variation to disease biology by testing whether genetically predicted gene expression is associated with a phenotype. PrediXcan tests genetically predicted expression against traits using reference transcriptome models, and S-PrediXcan extends this framework to genome-wide association study (GWAS) summary statistics ^4, 5^.

For drug repurposing, the direction of a TWAS or S-PrediXcan association can help prioritize therapeutic mechanisms. If increased genetically predicted expression of a gene is associated with increased disease risk, then an inhibitor or downregulator is directionally concordant with therapeutic benefit. If decreased genetically predicted expression is associated with increased disease risk, then an activator or upregulator is directionally concordant. Prior genetically informed drug-repurposing studies have used this logic to connect genetically predicted expression, drug targets, Mendelian randomization (MR), and electronic health record (EHR)-based validation and nominate candidate therapeutic inputs for multiple phenotypes ^6, 7^.

The bottleneck is that the step between a TWAS gene list and a directionally appropriate drug candidate is still commonly performed by manual curation. Investigators search drug-gene databases, read mechanistic literature, decide whether evidence suggests a drug inhibits, activates, upregulates, or downregulates the implicated gene, and then compares that direction with the TWAS effect estimate^7, 8^. This approach is biologically interpretable but does not scale easily to hundreds of genes and is difficult to reproduce across teams.

A related class of drug-repurposing approaches based on systems biology principles compares disease-level transcriptomic signatures with drug-induced gene network expression profiles, as introduced by the Connectivity Map and extended by later connectivity-mapping resources^9^. These approaches scale well and are valuable for whole-signature reversal, but they do not necessarily provide a gene-specific, literature-grounded explanation for a particular TWAS-nominated target. This can be an important element of subsequent efforts in animal models to expand indications and move to clinical trials.

We developed TRACE, a computational pipeline that automates the gene-by-gene curation step and implements the directionality logic that makes TWAS results useful for therapeutic hypothesis generation. The pipeline combines TWAS-derived effect direction, multi-database drug-gene retrieval, FDA approval filtering, PubMed evidence retrieval, and a fine-tuned biomedical language model that classifies drug-gene relationship support, mechanism, and direction from abstracts. We validate the system against three previously completed drug repurposing projects, two of which have been published, and apply it to endometriosis-associated TWAS genes as a proof-of-concept for precision-medicine drug repurposing and nominate several candidates with repurposing potential.

## 2. Methods

### 2.1. TRACE Pipeline overview

The pipeline accepts a gene symbol and a risk direction derived from a TWAS or PrediXcan effect estimate as input. It returns ranked FDA-approved drug-gene pairs classified as candidate therapeutic pairs, potential safety concerns, unclear-direction pairs requiring manual review, or drug-gene pairs without a specified disease-risk direction. The same code can be applied to any phenotype with a gene list and effect-size directions. Risk directions must be supplied by the user based on the sign of the effect estimate from their TWAS or S-PrediXcan output; a positive effect estimate corresponds to increased_gene_increases_risk and a negative effect estimate corresponds to decreased_gene_increases_risk. Once genes have been grouped by direction, the pipeline can be run on all genes in a group using the provided batch shell script, which loops over a user-supplied gene list and submits each gene with the appropriate direction flag. Users with large gene lists, for example several hundred genes from a GWAS or TWAS, do not need to run genes individually. Genes sharing the same direction can be batched together in a single command, and the two direction groups can be submitted sequentially or in parallel on a computing cluster. The pipeline source code and batch script examples are publicly available at https://github.com/otienoco/TRACE.

**Fig. 1.**
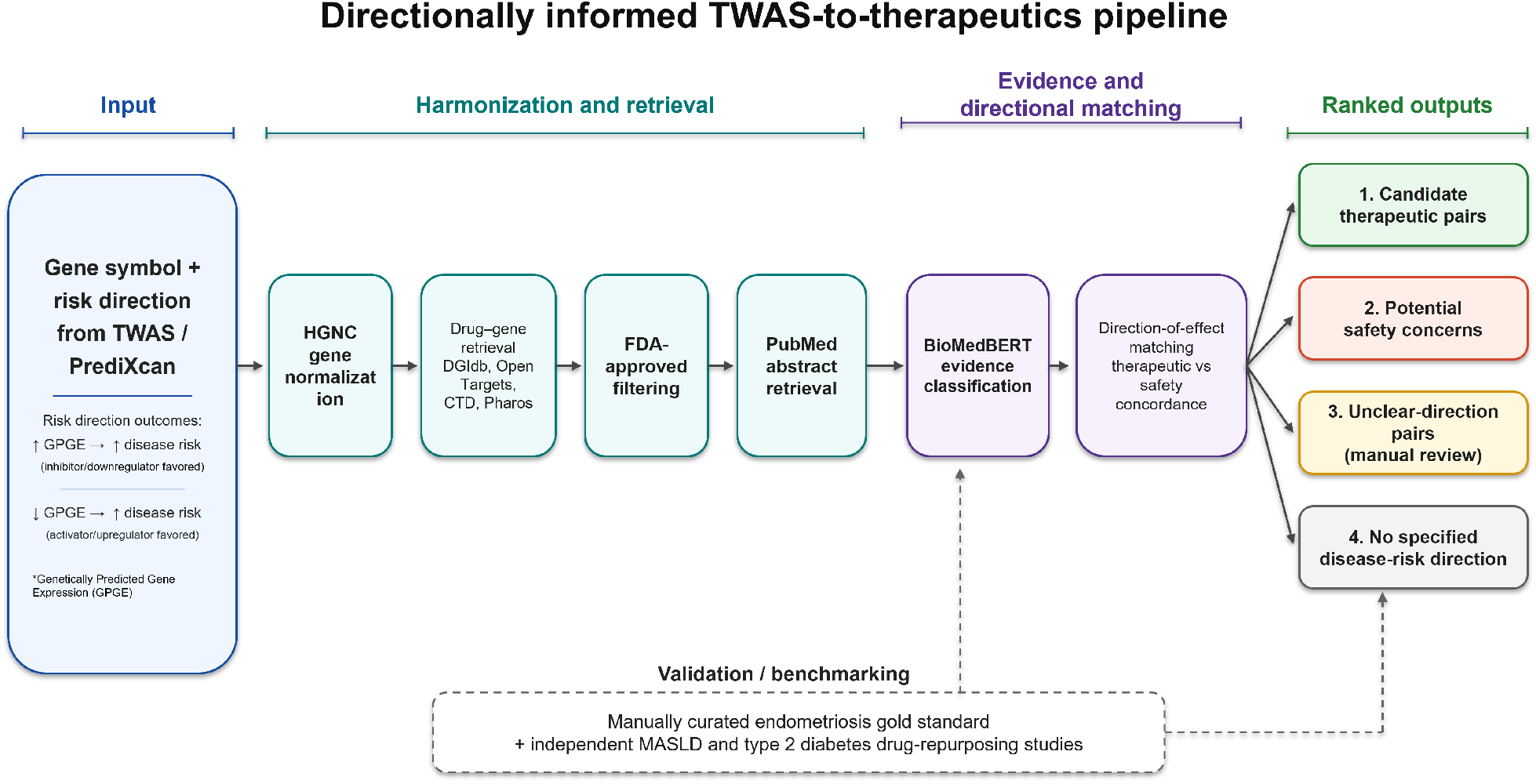
Overview of the directionally informed TWAS-to-therapeutics pipeline. A gene and TWAS effect direction are converted into FDA-approved drug-gene candidates, PubMed evidence, local BiomedBERT classifications, and prioritized therapeutic or safety hypotheses.

### 2.2. Gene normalization and drug candidate retrieval

Input gene symbols are resolved to official HGNC nomenclature through the HGNC REST Application Programming Interface (API), which supports queries against approved symbols, aliases, and previous symbols, allowing submitted symbols to be mapped to canonical HGNC identifiers before downstream database queries^10^.

Candidate drugs are retrieved from four drug-gene interaction resources. Drug Gene Interaction Database (DGIdb) 4.0 aggregates drug-gene interactions and druggable-gene information from publications, databases, and web resources^11^. Open Targets is an open-source platform designed to support systematic drug-target identification and prioritization^12^. CTD manually curates chemical-gene, chemical-disease, and related toxicogenomic relationships^13^. Pharos is the web interface for the NIH Illuminating the Druggable Genome Knowledge Management Center and provides integrated protein, target, and druggability information ^14^.

Drug candidates from the four resources are pooled and deduplicated by normalized drug name. Candidates are then filtered for confirmed United States FDA approval using the openFDA drug-labeling API, with RxNorm used to normalize brand names, generic names, and related clinical drug identifiers before matching across sources^15, 16^. Only confirmed FDA-approved compounds are advanced to literature retrieval and language-model classification steps.

### 2.3. PubMed evidence retrieval

For each FDA-approved drug-gene pair, PubMed abstracts are retrieved using the NCBI E-utilities API. Queries combine the official gene symbol and normalized drug name; PMIDs provided by source drug-gene databases are added as supplementary evidence when available. Up to seven abstracts per pair are retained by default, with each record storing the title, abstract text, PMID, and publication year. This value is configurable in config.py for users who wish to increase or decrease literature coverage per pair.

### 2.4. Evidence classification with a fine-tuned biomedical language model

Each abstract is classified on three axes: whether it supports a direct gene-level drug-gene relationship, the mechanism of the drug action on the gene, and the direction of the drug effect on gene expression or activity. Abstracts that document a relationship but do not state a clear direction are assigned an unclear-direction label rather than being forced into a mechanistic category.

The local classifier is built on BiomedBERT (microsoft/BiomedNLP-BiomedBERT-base-uncased-abstract-fulltext), a BERT-based biomedical language model pretrained on PubMed abstracts and PubMed Central full-text articles^17^. We added three classification heads to the [CLS] token representation for relationship detection, mechanism classification, and direction classification. The model was trained with a weighted cross-entropy loss summed across the three tasks. Training used the AdamW optimizer with a linear warmup schedule, a learning rate of 2×10^−5^, a batch size of eight, and five epochs on CPU.

During development, Claude API outputs were used to generate structured candidate labels and to identify disagreement cases for active learning. However, all validation and application results reported here were generated with the locally fine-tuned model only, with no external API calls during result generation. This design preserves transparency about model development while ensuring that the reported pipeline can run locally and without per-query API cost.

### 2.5. Training data and held-out model performance

Training data were built iteratively across four versions. Version 1 included 3,866 pipeline-derived examples. Version 2 added independent-disease validation examples. Version 3 incorporated the BioCreative VI ChemProt corpus, a manually annotated gold-standard corpus for chemical-protein relation extraction. ChemProt contains PubMed abstracts annotated for chemical and gene/protein entities and compound-protein relation types, including mechanistic relationships relevant to drug-target interpretation. This addition increased the number of expert-annotated training examples and exposed the classifier to curated chemical-protein relation labels rather than only pipeline-derived labels^18^. Version 4 added high-confidence active-learning disagreement cases reviewed during development.

**Table 1.**
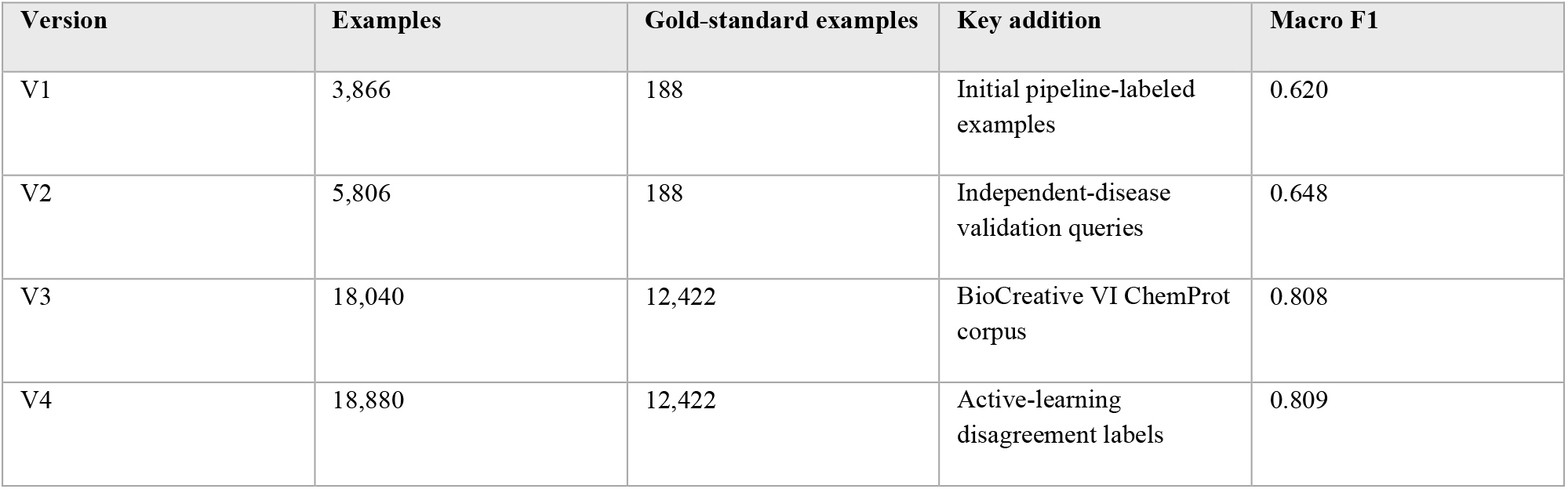
Training dataset versions and held-out macro F1.

| Version | Examples | Gold-standard examples | Key addition | Macro F1 |
| --- | --- | --- | --- | --- |
| V1 | 3,866 | 188 | Initial pipeline-labeled examples | 0.620 |
| V2 | 5,806 | 188 | Independent-disease validation queries | 0.648 |
| V3 | 18,040 | 12,422 | BioCreative VI ChemProt corpus | 0.808 |
| V4 | 18,880 | 12,422 | Active-learning disagreement labels | 0.809 |

The final model was evaluated on a held-out set of 2,833 examples. Relationship detection achieved 91% accuracy and 0.88 macro F1; mechanism classification achieved 84% accuracy and 0.78 macro F1; direction classification achieved 83% accuracy and 0.73 macro F1. Mean macro F1 across the three tasks was 0.809.

**Table 2.** Held-out performance of the final local classifier by task.

| Task | Accuracy | Macro F1 | Weighted F1 |
| --- | --- | --- | --- |
| Relationship detection | 91% | 0.88 | 0.91 |
| Mechanism classification | 84% | 0.78 | 0.84 |
| Direction classification | 83% | 0.73 | 0.83 |
| Overall mean | n/a | 0.809 | n/a |

### 2.6. Directionality classification and prioritization

Each gene is assigned a therapeutic direction from its TWAS effect estimate. A positive effect estimate, meaning increased genetically predicted expression is associated with increased disease risk, implies that inhibition or downregulation is the directionally concordant therapeutic hypothesis. A negative effect estimate implies that activation or upregulation is the directionally concordant therapeutic hypothesis. This follows the genetically informed drug-repurposing logic used in prior S-PrediXcan and MR-based studies^6, 7^.

Each drug-gene pair is then classified by comparing the literature-derived drug-effect direction with the gene-risk direction. A concordant pair is labeled candidate_therapeutic_pair; a discordant pair is labeled potential_safety_concern; a pair with documented relationship evidence but insufficient directionality is labeled unclear_direction_manual_review; and a pair with no disease-risk direction is labeled drug_gene_pair_only. Candidate pairs are ranked using a composite score incorporating evidence strength, number of supporting databases, classifier confidence, FDA approval, and supporting PubMed abstract count.

### 2.7. Validation strategy

We evaluated TRACE against three disease settings. Internal validation used 43 manually curated endometriosis drug-gene pairs across six TWAS-identified genes. External validation used published drug-gene pairs from a metabolic dysfunction-associated steatotic liver disease (MASLD) genetically informed repurposing study and from a type 2 diabetes EHR-validated genetically supported drug-repurposing pipeline^7, 8^. Drug aliases and salt forms were matched using a manually curated alias table, and all validation runs used the local classifier only.

## 3. Results

### 3.1. Internal validation of TRACE in endometriosis

Against the 43-pair manually curated endometriosis gold standard, the pipeline recovered 90.7% of pairs overall, including 88.9% of candidate therapeutic pairs and 92.0% of potential safety concern pairs. Exact-classification accuracy across the full gold standard was 46.5%, largely because many retrieved pairs were conservatively assigned to unclear_direction_manual_review rather than forced into candidate or safety categories when abstracts did not state a clear mechanism or direction.

The four unrecovered endometriosis pairs involved broad drug-class terms such as progestin that are not represented as specific FDA-approved compounds in the queried drug-gene databases. This failure mode reflects a database representation issue rather than a literature-classification failure.

**Table 3.**
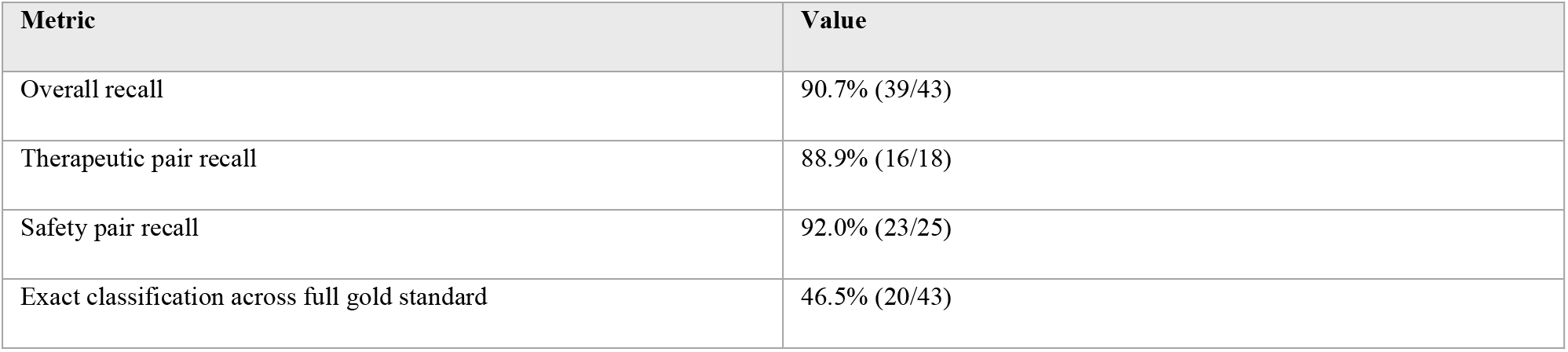
Internal validation results in the endometriosis gold standard.

### 3.2. External validation in MASLD and type 2 diabetes

In the MASLD validation set, the pipeline recovered 88.2% of published drug-gene pairs. In the type 2 diabetes validation set, raw recall was 65.0%; however, six of seven unrecovered pairs were explained by database coverage or mapping limitations, including withdrawn drugs, literature-supported pairs absent from the four queried databases, and calcium-channel subunit mapping inconsistencies. Restricting the denominator to pairs present in at least one queried database yielded 92.9% adjusted recall.

**Table 4.** External validation across two independent disease applications.

| Disease | Source | Pairs | Recall | Adjusted recall |
| --- | --- | --- | --- | --- |
| MASLD | Seagle et al., 2025 | 34 | 88.2% (30/34) | n/a |
|  | (PMID: 40780050) |  |  |  |
| Type 2 diabetes | Shuey et al., 2023 (PMID: 37399599) | 20 | 65.0% (13/20) | 92.9% (13/14) |

Across the three disease settings, the pipeline recovered 88-93% of curated or database-findable pairs, suggesting that the main remaining limitation is drug-gene database coverage rather than PubMed retrieval or local language-model classification.

### 3.3. Application to endometriosis-associated TWAS genes

We applied TRACE to 99 endometriosis-associated TWAS genes identified through S-PrediXcan analysis across 49 human tissues from the Genotype-Tissue Expression (GTEx) project version 8. Genes were retained if they passed a false discovery rate threshold of 0.05 and a colocalization posterior probability threshold of 0.60, yielding 49 genes requiring an inhibitory or downregulating therapeutic effect, 45 genes requiring an activating or upregulating effect, and five genes with ambiguous naming. Of the 99 genes, 75 had at least one FDA-approved drug interaction across the four queried databases. Across those druggable genes, the pipeline identified 1,089 FDA-approved drug-gene pairs, including 32 candidate therapeutic pairs and 77 potential safety concerns. The full 99-gene analysis was completed in approximately two hours on a standard high-performance computing cluster node using the local classifier, at no per-query API cost, with individual gene queries requiring approximately 10 to 20 minutes depending on the number of FDA-approved candidates retrieved.

The pipeline recovered 39 of 43 manually curated endometriosis validation pairs when run with the local model, matching the recall obtained during development with API-assisted classification. This indicates that the local classifier reproduced the retrieval and prioritization performance needed for the application analysis while avoiding external API use during final result generation.

Beyond the manually curated genes, the pipeline surfaced 27 additional candidate therapeutic pairs across 21 genes. For *GNRH1*, the pipeline independently identified leuprolide acetate, a GnRH agonist that has randomized-trial evidence supporting its efficacy in endometriosis-associated pain and disease suppression^19-21^. We treat this result as an internal positive control showing that the pipeline can rediscover a clinically established endometriosis therapy from automated evidence retrieval and classification alone.

**Table 5.**
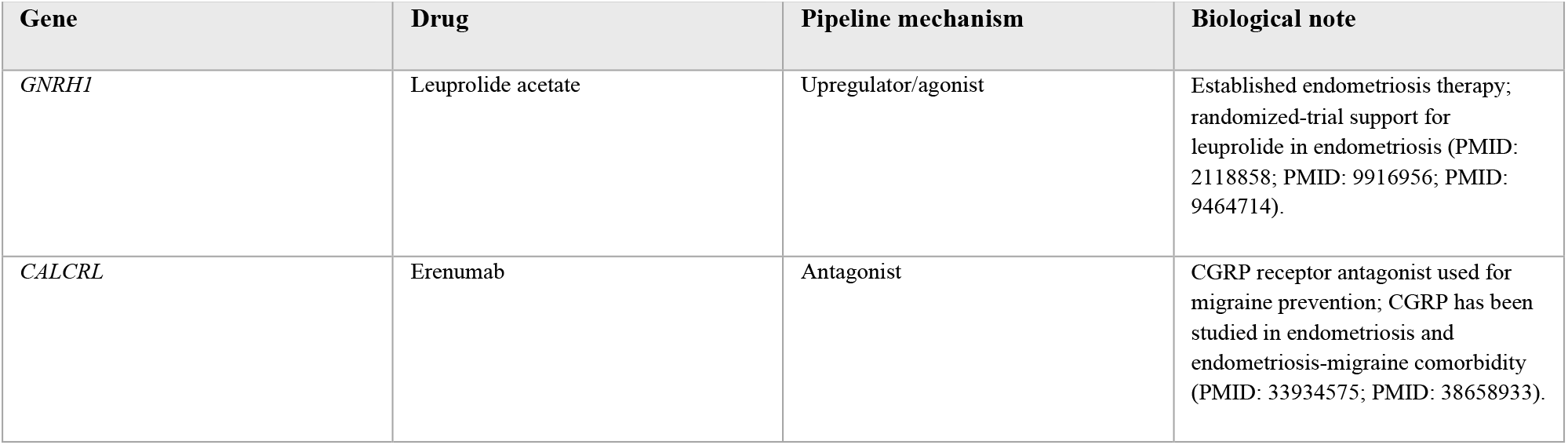
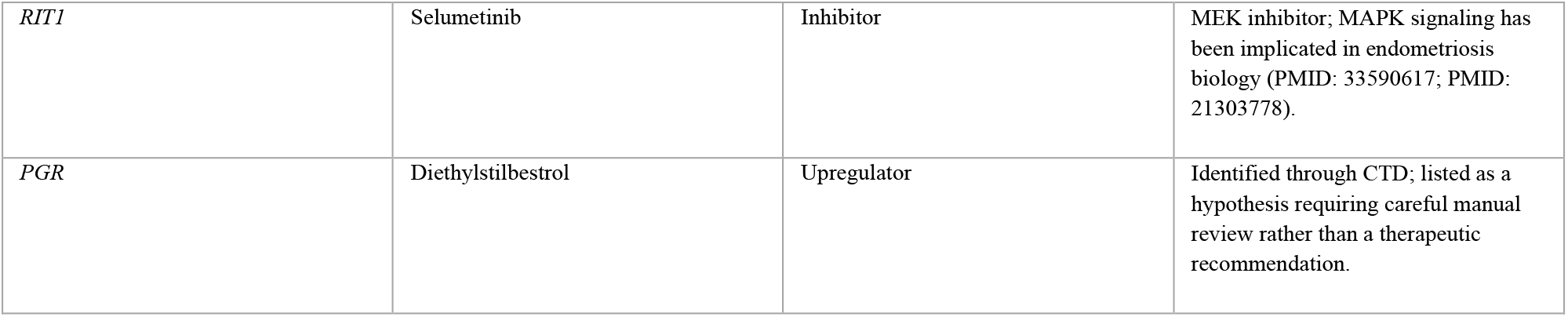
Selected candidate therapeutic pairs identified by the pipeline in the endometriosis application.

## 4. Discussion

We developed and validated TRACE, a scalable computational pipeline that automates a key bottleneck in genetically informed drug repurposing: converting TWAS-nominated genes and effect directions into literature-supported, directionally classified drug-gene hypotheses. This method differs from prior manual genetically informed repurposing workflows by extracting drug-gene direction from PubMed evidence at the pair level, applying the same logic uniformly across large gene lists, and replacing API-assisted development labels with a local fine-tuned biomedical language model for final validation and application, creating a scalable, reproducible drug repurposing pipeline.

The main strength of the framework is that it preserves the interpretability of gene-specific curation while improving scalability and reproducibility. Rather than assigning a drug direction from a broad therapeutic class or clinical indication, the model evaluates abstracts for evidence about the specific drug-gene pair. This is important because the same compound can have different mechanisms at different targets or in different biological contexts.

The validation results support the use of the pipeline as a high-recall prioritization and triage tool. In endometriosis, MASLD, and type 2 diabetes, the pipeline recovered most curated or database-findable pairs. When it could not assign a confident direction, it frequently used the unclear_direction_manual_review category rather than forcing a potentially incorrect candidate or safety label. This conservative behavior reduces overclaiming and makes the output more appropriate for translational hypothesis generation.

The endometriosis application illustrates how the system can expand manual review. A human team might reasonably focus on a few high-priority genes, but the pipeline applies the same process to all genes with assigned risk direction, producing candidate pairs, safety concerns, and manual-review cases across the full TWAS gene set. The independent recovery of leuprolide acetate provides a positive control^19, 21, 22^, while candidates such as erenumab and selumetinib illustrate how the method can propose biologically plausible hypotheses that require downstream validation.

The pipeline does not establish therapeutic efficacy. It produces prioritized hypotheses that should be followed by the same validation steps used in prior genetically informed repurposing work, including MR with gene-expression or protein-level proxies, EHR-based validation when appropriate, and experimental assays or structural modeling when biologically informative^6-8^.

This study also has limitations. First, the pipeline depends on the coverage and quality of source drug-gene databases; pairs absent from DGIdb, Open Targets, CTD, and Pharos cannot be recovered unless future versions add additional resources. Second, restricting the search to FDA-approved drugs increases immediate translational relevance but excludes investigational drugs and compounds approved only outside the United States. Future versions of this package will include an option to label by FDA status rather than filter. Third, some mechanism classes remain underrepresented in the training data, limiting classifier performance for rare classes. Fourth, the unclear-direction category was common, reflecting the fact that abstracts often document drug-gene relationships without stating a direct expression or activity direction.

Future work will focus on expanding active-learning labels across additional diseases and incorporating additional drug-target and pharmacogenomic resources. Because the current input is simply a gene symbol and effect-size direction, TRACE is readily applicable to any complex trait or disease with TWAS, PrediXcan, or mapped gene results. As GWAS and TWAS resources continue to grow across diverse ancestries and tissues, tools that can rapidly and reproducibly translate genetic discovery into directionally informed therapeutic hypotheses will become increasingly valuable. TRACE provides that bridge, offering a scalable, cost-free, and literature-grounded approach to drug repurposing that can accelerate the path from genetic association to prioritized candidates for experimental and clinical follow-up.

## Data Availability

All data produced in the present work are contained in the manuscript. The TRACE pipeline code is publicly available at https://github.com/otienoco/TRACE and the fine-tuned model weights are available at https://huggingface.co/otienoco/TRACE-classifier.

https://huggingface.co/otienoco/TRACE-classifier

## Acknowledgments

Research reported in this publication was supported by the Eunice Kennedy Shriver National Institute of Child Health and Human Development of the National Institutes of Health under award number R01HD110567. H.M.S. was supported by American Heart Association grant 26PRE1550935 and National Institutes of Health grant TL1TR002244. C.O.O. was supported by National Institutes of Health grant TL1TR002244. A.T.A. was supported by National Institutes of Health grant T32 CA160056. Preprint of an article submitted for consideration in Pacific Symposium on Biocomputing © 2027 World Scientific Publishing Co., Singapore, http://psb.stanford.edu/. The authors wish to dedicate this work to the memory of Dr. Ephraim Otieno, MD, whose thoughtful curiosity and encouragement during the development of this work are deeply appreciated.

## Data Availability

The TRACE pipeline source code is publicly available at https://github.com/otienoco/TRACE. The fine-tuned BiomedBERT classifier weights are available at https://huggingface.co/otienoco/TRACE-classifier and are downloaded automatically on first run when using the local classifier.

## LLM Disclosure

The Claude Application Programming Interface (API) (Anthropic) was used during pipeline development as an evidence classifier and as one source of labels for active-learning disagreement cases, as described in Methods. All validation and endometriosis application results reported in this manuscript were generated using the locally fine-tuned model only, with no Claude API involvement during result generation. Claude was also used as a coding assistant during pipeline development.

